# Microvascular insulin resistance associates with enhanced muscle glucose disposal in CD36 deficiency

**DOI:** 10.1101/2024.02.16.24302950

**Authors:** Cyndya Shibao, Vivek S. Peche, Ian M. Williams, Dmitri Samovski, Terri A. Pietka, Naji N. Abumrad, Eric Gamazon, Ira J. Goldberg, David Wasserman, Nada A. Abumrad

**Affiliations:** Department of Medicine, Division of Clinical Pharmacology, Vanderbilt University Medical Center, Nashville TN; Department of Medicine, Division of Nutritional Sciences and Obesity Research, Washington University School of Medicine, St. Louis, MO; Department of Molecular Physiology and Biophysics, Vanderbilt University Medical Center, Nashville TN; Department of Surgery, Vanderbilt University Medical Center, Nashville TN; Department of Medicine, Division of Genetic Medicine, Vanderbilt University, Nashville, TN; Department of Medicine, Division of Endocrinology, Diabetes and Metabolism, New York University Grossman School of Medicine, New York, NY; Department of Cell Biology & Physiology, Washington University School of Medicine, St. Louis, MO

**Keywords:** Endothelial function, microvascular circulation, nitric oxide, African Americans

## Abstract

Dysfunction of endothelial insulin delivery to muscle associates with insulin resistance. CD36, a fatty acid transporter and modulator of insulin signaling is abundant in endothelial cells, especially in capillaries. Humans with inherited 50% reduction in CD36 expression have endothelial dysfunction but whether it is associated with insulin resistance is unclear. Using hyperinsulinemic/euglycemic clamps in *Cd36^-/-^* and wildtype mice, and in 50% CD36 deficient humans and matched controls we found that *Cd36^-/-^* mice have enhanced systemic glucose disposal despite unaltered transendothelial insulin transfer and reductions in microvascular perfusion and blood vessel compliance. Partially CD36 deficient humans also have better glucose disposal than controls with no capillary recruitment by insulin. CD36 knockdown in primary human-derived microvascular cells impairs insulin action on AKT, endothelial nitric oxide synthase, and nitric oxide release. Thus, insulin resistance of microvascular function in CD36 deficiency paradoxically associates with increased glucose utilization, likely through a remodeling of muscle gene expression.

## Introduction

The membrane fatty acid (FA) transporter CD36 was initially identified as an insulin resistance gene when CD36 sequence variants were found in the insulin-resistant spontaneously hypertensive rat (SHR) a model of type 2 diabetes (T2D). Transgenic CD36 expression ameliorated the defect in FA metabolism and insulin resistance of the SHR (1, 2). In humans a C/T single nucleotide polymorphism (SNP rs1527479) in the CD36 promoter associates with insulin resistance and T2D (3), a rare nonsense mutation (rs56381858) in a Caucasian pedigree is linked to autosomal dominant T2D (4). Our genome-wide association analysis of available data from the Vanderbilt BioVu patient biobank and the MAGIC Consortium show that reduced CD36 expression in muscle and heart, and a SNP (rs17236824) within 1 MB of the transcription start site, associated with T2D renal, ophthalmic, and neurological complications (5). In addition, a common CD36 haplotype is associated with T2D-linked coronary heart disease (6). The mechanisms underlying the T2D-related metabolic alterations that are associated with CD36 variants remain unclear.

Complete CD36 deficiency has a prevalence of 0.3–11% with higher incidences in populations of Asian and African descent (7, 8). CD36 coding SNP rs3211938 (G/T) is the major cause of deficiency in African Americans and carriers of the minor G allele (frequency 20-25%) have 50% reduction of CD36 levels (9, 10). Although deficiency is low in Caucasians (0.3%) SNPs that reduce CD36 levels are common (11). Information on whether insulin sensitivity is altered in individuals with partial or total CD36 deficiency is limited. Only few studies evaluated whole-body glucose disposal in small cohorts of these subjects and reported conflicting results (12-14). Studies in mice suggest a complex relationship between CD36 expression and insulin sensitivity. *Cd36**^-/-^*** mice have enhanced systemic glucose disposal, consistent with better muscle insulin sensitivity (15), however this enhancement is not recapitulated by CD36 deletion in muscle (5). CD36 knockdown in cultured human myotubes impairs insulin signaling, as CD36 promotes tyrosine phosphorylation of the insulin receptor (IR) by Fyn kinase and enhances IR signaling to AKT (5). Surprisingly, in contrast to the effect of muscle CD36 deletion, endothelial cell (EC) specific deletion enhances systemic glucose disposal (16) mimicking the *Cd36**^-/-^*** mouse phenotype (16). These data can be interpreted to indicate that muscle glucose disposal is critically influenced by endothelial CD36. In the present study we interrogated some of the underlying mechanisms and their potential relevance to humans.

The vasculature is the most important regulator of nutrient exchange and dysfunction of microvessels can alter muscle access to circulating insulin and glucose (17, 18). Strong evidence supports importance of vascular insulin action in regulating muscle glucose disposal (19, 20). For example the endothelial dysfunction that associates with obesity causes a delay in insulin action to increase peripheral blood flow, and correlates with reduced whole body glucose uptake (21). We previously described reduced flow mediated dilation of the brachial artery and low cGMP levels in subjects carrying the G-allele of CD36 coding SNP rs3211938, suggesting reduced nitric oxide (NO) bioavailability (9). In this study we used metabolic insulin clamps to assess insulin sensitivity of glucose disposal in *Cd36**^-/-^*** mice, and in 50% CD36 deficient human carriers of the G-allele of SNP rs3211938, we examined the relationship between microvascular function, transendothelial insulin flux and muscle glucose uptake.

## Methods

### Mice Experimental Protocols

Mice studies used age-matched 3-4 months old wild-type (WT) and *Cd36^−/−^*mice on the C57BL/6 background. Male and female mice were used except when indicated. Experimental protocols were approved by the Animal Studies Committees of Washington University or Vanderbilt University.

#### Vessel Compliance

Ascending aortas and left carotid arteries (from transverse aorta to 6 mm up the common carotid) were dissected from euthanized WT and *Cd36*^-/-^ mice as previously described (22, 23). Briefly, vessels mounted in physiological saline on a pressure arteriograph (Danish Myotechnology, Copenhagen), were pressurized and longitudinally stretched to *in vivo* length three times before data capture. Vessel diameter recordings used a transillumination microscope connected to a camera and a computerized measurement system (Myoview, Danish Myotechnology). Intravascular pressure was increased from 0 to 175 mmHg in 25-mmHg steps and the vessel’s outer diameter (OD) was measured at each step.

#### Muscle insulin sensitivity in vivo

5 hour fasted mice were injected with intraperitoneal insulin (0.75 units/kg) and tissues harvested 15 min later were quickly frozen in liquid nitrogen and kept at −80°C until used for western blots as previously described (5). Antibodies used: anti-mouse CD36 (R&D Systems, AF1955), anti-pS473-Akt (Cell Signal Technology, 4060), anti-pT308-Akt (Cell Signal Technology, 13038) and total Akt (Cell Signal Technology, 4691).

#### Transcapillary Insulin Flux

Intravital microscopy (IVM) of the exposed gastrocnemius was used to visualize transcapillary insulin flux in WT and *Cd36^-/-^* mice. Insulin was conjugated to Alexa Fluor 647 and the probe tested to ensure purity and bioactivity comparable to unconjugated insulin as previously described (24). Prior to IVM, the probe was mixed vigorously in saline, sonicated 1 hour, centrifuged (20 min, 13000 rpm) to remove insoluble material and microscopically inspected for absence of aggregates (24). The mice were anesthetized with ketamine/xylazine/acepromazine (7.9/1.6/0.2 mg/kg) via an indwelling venous catheter, placed on a heated pad. Skin of the shaved lower leg was removed to expose the lateral gastrocnemius and peel off its fascia avoiding perturbation of muscle fibers and blood vessels (24). For imaging the mouse was transferred to the custom stage mount and its body temperature kept at 37°C through a feedback control system connected to an electric blanket (Harvard Apparatus). To keep hydration and physiological stability of the gastrocnemius it was continuously irrigated with bicarbonate-buffered physiological saline solution (PSS); in mmol/L 132 NaCl, 4.7 KCl, 2 MgSO_4_, 1.2 CaCl_2_, and 18 NaHCO_3_. PSS kept at 37°C was bubbled with 95%N_2_/5%CO_2_ to maintain a 7.4 pH and continuously circulated over the tissue using a peristaltic pump (inflow) and a vacuum trap (outflow) (24).

Quality of the gastrocnemius preparation was visualized using tissue autofluorescence with light from a 120W mercury arc lamp (XCite^®^ 120) sent through a bandpass filter (Zeiss Filter Set 10; Excitation: 450-490nm, FT: 510nm, Emission: 515-565nm) onto the exposed region. To visualize blood vessels the mouse was infused with 50 μg of 2 MDa rhodamine-dextran (rho-dex) and the filter set switched (Zeiss Filter Set 43 HE; Excitation: 550/25nm, FT:570nm, Emission: BP 605/70). The field of view was selected from a capillary bed stemming from the external sural artery based on 1) presence of a sufficient number of clearly visible capillaries; 2) absence of large vessels in nearby area; and 3) absence of capillaries immediately adjacent to one another.

Intravital microscopy used an inverted Zeiss LSM 780 microscope (Zen software) with a 20X 0.8NA Plan-Apochromat air objective. For imaging transendothelial molecular flux (ins-647, BSA-647, dextrans), rho-dex fluorescence was excited with light from a 561 nm solid-state laser and detected on a multichannel PMT. Near-infrared fluorophores were excited by a helium-neon 633nm laser and emitted light detected with a gallium arsenide phosphide semiconductor. For both rho-dex and near-infrared fluorophores, excitation and emission light were reflected with an MBS 488/561/633 dichroic mirror and imaging used two-channel sequential excitation and detection to avoid bleed through, in an optical section of 8 μm (+/-4μm about the focal plane). The 8-bit intensity, 1024x1024 pixel images were acquired with unidirectional scanning. For each time point, a 4-“slice” z-stack was acquired in each channel using a 4μm step size to avoid aliasing. Photomultiplier tube settings were adjusted to maximize image dynamic range and kept constant for each experiment for quantitative comparisons. The imaging region was selected, a background image (t=0) was acquired then probe (rho-dex and ins-647) was injected through the venous catheter and images acquired every minute at t=1-10, at 12.5 and 15 min, post probe injection (24).

#### Hyperinsulinemic-euglycemic Insulin Clamp

Catheters were implanted under isoflurane anesthesia in the carotid artery for sampling and jugular vein for infusion a week before performing clamps (25) to allow the mice to regain weight lost due to surgery. The mice were fasted for 5h before the clamp and not restrained or handled during the experiments (26). [3-^3^H]glucose was primed and continuously infused from *t*=-90min to *t*=0min (0.04 µCi/min). The insulin clamp was initiated at *t*=0 min with a continuous insulin infusion (4 mU·kg^-1^·min^-1^) and a variable glucose infusion rate (GIR), both maintained until t=155 min. Infused glucose had 0.06 µCi/µL [3-^3^H]-glucose to minimize changes in plasma [3-^3^H]-glucose specific activity. Arterial glucose was measured every 10 min and GIR adjusted to maintain euglycemia. Erythrocytes were infused to compensate for withdrawn blood. [3-^3^H]-glucose kinetics were determined at -15 min and -5 min (basal period) and every 10 min between 80-120 min to assess whole-body glucose appearance (R_a_), disappearance (R_d_), and endogenous glucose production (endoR_a_). For tissue-specific index of glucose uptake (R_g_) intravenous 13 µCi 2-[^14^C]-deoxyglucose ([^14^C]2DG) was given at 120 min and blood samples collected at 122, 125, 135, 145 and 155 min to assess [^14^C]2DG plasma disappearance. Plasma [3-^3^H]-glucose and [^14^C]2DG, and tissue [^14^C]2DG-phosphate were measured as previously (27). At 155 min, the mice were anesthetized, and harvested tissues freeze clamped. Protocol details are available online at the Vanderbilt Mouse Metabolic Phenotyping Center (www.vmmpc.org).

#### RNAseq and Microarray

For RNA seq human microvascular cells were treated with either control or CD36 siRNA and 72 h later total RNA isolated (TRIzol Reagent, ThermoFisher Scientific), and cleaned (RNeasy, Qiagen). Samples were submitted to the Washington University Genome Technology Access Center (GTAC) for library preparation, sequencing and read alignment. Normalization and differential gene expression used the integrated Differential Expression and Pathway Analysis (iDEP) web platform (28). For microarrays total RNA (TRIzol) isolated from heart of 4h fasted mice was analyzed using Whole Mouse Genome Oligo Microarray (Agilent Technology, Santa Clara, CA) at the Washington University Functional Genomics Core of the Digestive Disease Research Center.

### Human Study Protocols

#### Subjects

Healthy unrelated African American men and women, between the ages of 18-50 years old were recruited and genotyped for the CD36 coding single nucleotide polymorphism (SNP) rs3211938 (G/T). This SNP, which reflects positive selection pressure, is almost exclusive to populations of African ancestry (where G-allele incidence is 20-25%) (9). The SNP inserts a stop codon resulting in a truncated protein that is degraded. Partial CD36 protein deficiency is observed in subjects carrying the G allele and complete deficiency is observed in subjects who are homozygous (G/G) (10). Subjects were excluded from participating in the study for the following: BMI>40 kg/m^2^, type 1 or 2 diabetes mellitus, cardiovascular disease including hypertension, impaired renal or liver function, use of nitrates, or systemic glucocorticoid therapy. Participants reported to the Vanderbilt Clinical Research Center (CRC) for pre-screening and blood collection. DNA was extracted, genotyped for rs3211938, and G-allele carriers and noncarriers invited to a screening visit for a medical history, physical exam, and laboratory analyses (blood cell count, metabolic panel, pregnancy test).

Genomic DNA was extracted from peripheral blood with a salting-out precipitation (Gentra Puregene) and CD36 SNP rs3211938 detected using a predesigned TaqMan SNP genotyping assay (Applied Biosystems) on a 7500 Fast (Applied Biosystems) instrument. Previous studies have identified that the rs3211938 G allele reduces CD36 expression by 50% (9, 10). All human assessments and studies were performed at the Vanderbilt CRC at 8 am in a quiet, temperature-controlled room (22 to 23 ^◦^C). Participants were asked not to exercise or drink alcohol for at least 24 hours before the study and were contacted by the research nurse to remind them to fast the night before. All participants underwent two visits at least 2 weeks apart. All studies adhered to principles of the Declaration of Helsinki and Title 45 of the US Code of Federal Regulations (Part 46, Protection of Human Subjects). Studies were approved by the Vanderbilt Institutional Review Board and conducted in accordance with institutional guidelines. All subjects provided informed consent. The study was included in clinicaltrials.gov, identifier: NCT03012386

#### Hyperinsulinemic-euglycemic (Insulin) Clamp

During each visit, two intravenous (IV) catheters were placed in the antecubital fossa of each arm. One of the catheters was used for drug infusion, the other one for blood collection. A linear-array transducer connected to an ultrasound was placed on the brachioradialis muscle of the arm not used for infusion. The transducer was used to measure changes in microvascular circulation with contrast-enhanced ultrasonography (CEU) and remained in the same position throughout the study.

On the first study day (saline day), participants received 0.9% standard saline infusion (45ml/h) for 6 hours, and during the last three hours, a hyperinsulinemic-euglycemic (HIE) clamp. Insulin was infused at a rate of 80 mU·m^-2^·min^-1^ for 5 min followed by 40 mU·m^-2^·min^-1^ for the remainder of the study. Plasma glucose samples were obtained every 5 minutes throughout the clamp and 5% glucose was infused at a variable rate to maintain plasma glucose between 90 to 95 mg/dl. Blood samples were collected for assay of plasma insulin.

The second study day was ∼4 weeks later. Participants returned to the CRC and instead of saline received a 6-hour IV infusion of 20% Intralipid^®^ (Baxter Healthcare Crop. Glendale, CA) at a rate of 45 ml/h and heparin (200 units prime followed by 200 units/hr.). Heparin was used to activate endothelial lipoprotein lipase and accelerate lipolysis of triglycerides into fatty acids. The HIE clamp was conducting during the last three hours of the study.

In both day visits, CEU measurements were obtained 3 hours after initiation of infusions (saline or Intralipid^®^ and heparin) during the insulin clamp. Intermittent blood pressures and ECG were measured using the VITAL-GUARD 450c monitor (Ivy Biomedical Systems, Branford, CT). Vanderbilt’s Investigational Pharmacy conducted drug preparation, storage, and dispensing logs.

#### Assessment of microvascular blood volume index

Contrast-enhanced ultrasound imaging used a linear-array transducer connected to an ultrasound (L9-3 mm transducer, iU22; Phillips). Real-time imaging used low (0.08) and high (1.2) mechanical index. The contrast microbubbles (Definity, Bristol-Myers Squibb) were infused at 1.5 ml/min for 10 min. At steady state (∼4 min), the 1.2 mechanical index destroyed the microbubbles at the start of video recording. Switch to the low index (0.08) made microbubbles resonate allowing real-time recording of vascular replenishment. Local temperature was measured with a laser Non-Contact Infrared Skin Thermometer. Data were analyzed using QLAB software.

#### Clinical Chemistry

Blood collected in chilled EDTA tubes was immediately centrifuged and separated plasma stored at −80°C. For serum, blood clotted at room temperature for 20 minutes was centrifuged and removed serum stored at −80°C. Plasma glucose was measured with a glucose analyzer (YSI Life Sciences, Yellow Springs, OH). Plasma insulin concentrations were determined by radioimmunoassay (Millipore, St. Charles, MO).

### Human microvascular cells (hMEC) protocols

#### CD36 knockdown, Caveolin 1-eNOS interaction

Human derived primary dermal microvascular cells, hMEC, (Lonza bioscience) were cultured in EGM-2 MV endothelial media (Lonza) and passaged less than 5 times. Cells were treated with siRNA against CD36 (4392420, assay ID 2646, ThermoFisher Scientific) or with control siRNA for 72 hours, as previously described (28). The hMEC were serum starved for 4 h before adding human insulin (100nM, Sigma) for the indicated times. For western blots, cells were lysed 20 min in ice-cold RIPA buffer (20 mM Tris-HCl, pH 7.5, 150 mM NaCl, 1mM Na2EDTA, 1 mM EGTA, 1% NP-40, 1% sodium deoxycholate, 2.5 mM sodium pyrophosphate, 1 mM beta-glycerophosphate, 1 mM Na3VO4, 1µg/mL leupeptin, 1mM PMSF, and 1μg/mL protease inhibitor mix). Cleared lysates (10,000 x g, 10 min) were assayed for protein content (DC Protein Assay, Bio-Rad). Separated proteins (30 µg, 4%–20% gradient gels, ThermoFisher Scientific) were transferred to polyvinylidene membranes (Immobilon Fl, Millipore), blocked (1X TBS, 0.25% fish gelatin, 0.01 % Na-azide, 0.05% Tween-20) and primary antibodies added overnight at 4°C. Washed membranes were treated 1h with infrared dye–labeled secondary antibodies (RT, LI-COR Biosciences) and imaged (LI-COR Odyssey Infrared). Primary antibodies used: anti-human CD36 (R&D Systems, AF2519), anti-pS473-Akt (Cell Signal Technology, 4060), total Akt (Cell Signal Technology, 4691), pS1177-eNOS (Cell Signal Technology, 9571), total-eNOS (Cell Signal Technology, 32027) and beta-Actin (Santa Cruz Biotechnology, sc-47778).

For immunoprecipitation (IP) 4h serum-starved hMECs were treated with insulin (100nM 10min, 37°C) then washed cells were scraped into 1 ml IP buffer (0.1 M NaCl, 0.3 M Sucrose, 30mM MgCl2, 10 mM PIPES, 0. 5mM EDTA, 0.1% Nonidet P-40, protease and phosphatase inhibitors and 0.5 mM pervanadate) kept on ice 20 min, then lysed by passage 5 times through a 25-gauge needle. Aliquots (50 µg protein) of clarified (11,000 x g, 10 min) lysates were incubated 5h with Cav-1 antibody (Cell Signaling Technology, 3238) coupled to protein G magnetic beads (Dynabeads, ThermoFisher). Rabbit IgG-coupled beads were controls. Immune complexes were separated magnetically, washed (IP buffer) and boiled in 50µL of 2X SDS-Sample buffer prior to SDS-PAGE (4-12%).

#### Nitric Oxide measurement

The Nitrate/Nitrite Colorimetric Assay (Cayman Chemicals, 780001) was used. Human insulin (Sigma, I9278) was added (100nM, 10 min) to 4 h serum starved cells and media collected for NO measurement as per the manufacturer’s protocol.

### Biostatistics

Data are presented as means ± standard errors of the mean (SEM). Data were log-transformed prior to statistical testing when not normally distributed by the Shapiro–Wilks test. Summary data were analyzed by a paired, parametric, two-tailed Student t test. P values of <0.05 were considered significant (****P ≤ 0.0001, ***P ≤ 0.001, **P ≤ 0.01, *P ≤ 0.05, ns P > 0.05). All analyses used Prism 10.0 (GraphPad Software, La Jolla, CA).

## Results

### CD36_-/-_ mice have enhanced systemic glucose disposal and muscle insulin signaling

Hyperinsulinemic glucose clamps were performed in mice following a 5 h fast and a period of basal sampling. *Cd36^-/-^* mice have slightly smaller body weight than WT mice (**Fig. 1A**), on average around 8% with similar lean body mass (29). Under basal conditions, plasma insulin was similar for WT and *Cd36^-/-^* mice but the latter group had lower arterial glucose, in line with earlier studies (15, 16). Clamping glucose during equivalent hyperinsulinemia required a significantly higher GIR in *Cd36^-/-^* than in WT (**Fig. 1B, C, D**). Whole body rate of disposal (Rd) was increased during the clamp in *Cd36^-/-^* compared to WT mice (**Fig. 1E**) and the *Cd36^-/-^* mice displayed greater % suppression of endogenous rate of appearance (Ra) during the insulin clamp (85% versus 59%) (**Fig. 1F**). The delta and percent suppression of Ra were both significant. The absolute Ra was close to being significant <0.069 and lower Ra contributes to GIR. The estimated contribution of Rd and Ra to the greater GIR was respectively 2:1.

**Figure 1.**
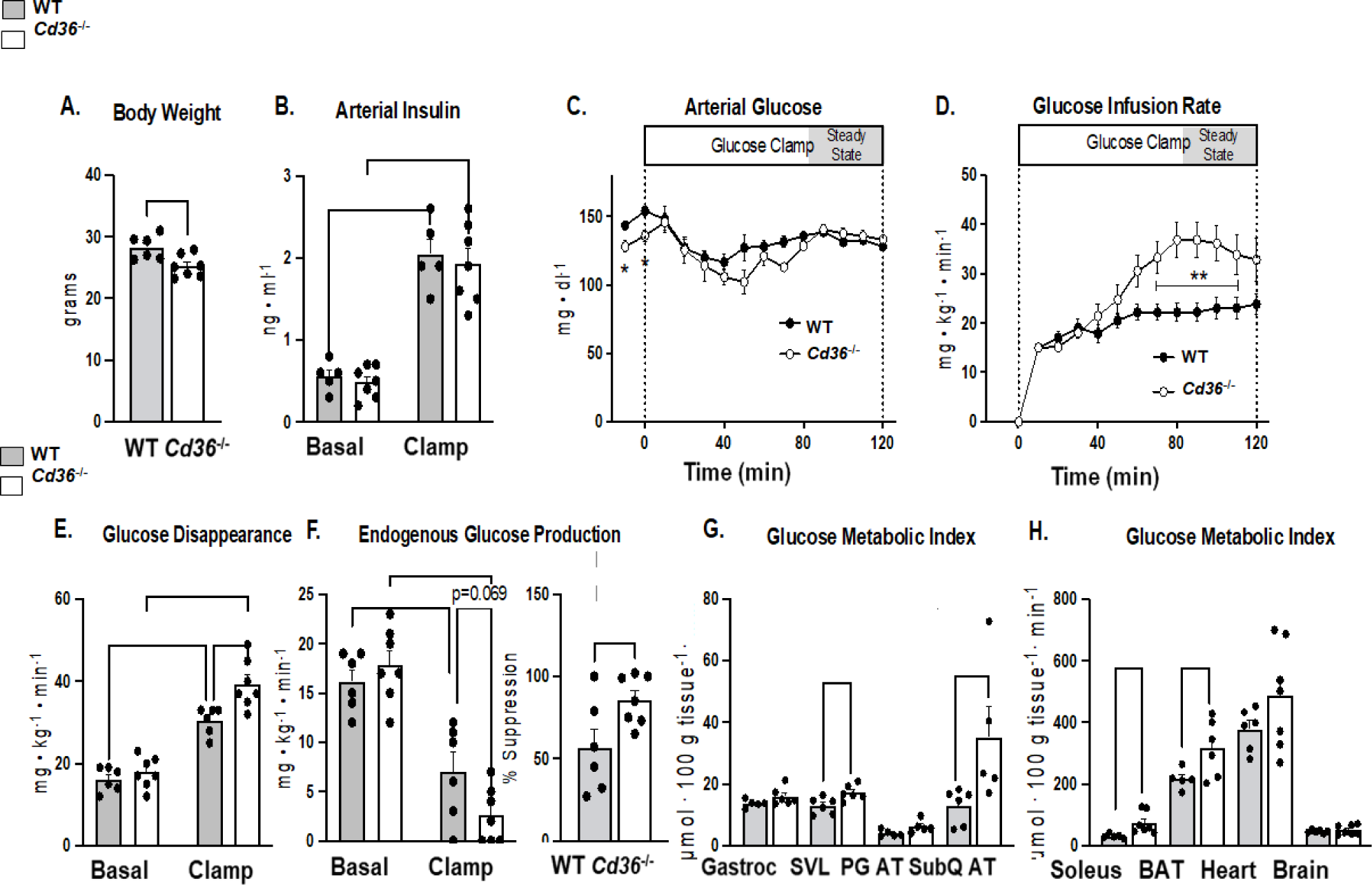
CD36 deletion in mice improves insulin sensitivity and the glucose metabolic index. Hyperinsulinemic-euglycemic clamps were performed in awake, non- restrained wildtype (WT) and *Cd36****-/-*** mice. **A.** Body weight. **B**. Fasting and clamp insulin was similar in both genotypes. **C**. Reduced fasting glucose in *Cd36****-/****-* but similar blood glucose levels in WT and *Cd36****-/-*** mice during the clamp. **D**. Higher glucose infusion rates were needed to maintain euglycemia in *Cd36****-/-*** mice compared to WT. **E**. Glucose disappearance rates are higher in *Cd36****-/-*** mice during the clamp. **F**. Insulin more effectively suppresses endogenous glucose production in *Cd36****-/-*** mice. **G**-**H**. Glucose metabolic index in superficial vastus muscle (SVL), soleus, subcutaneous (SubQ AT), and brown adipose tissue (BAT) in *Cd36****-/-*** mice as compared to WT mice. Gastrocnemius (gastroc), peri- gonadal adipose tissue (PG AT), and heart glucose metabolic index were elevated but differences did not reach significance. As expected, brain glucose metabolic index was equivalent in WT and *Cd36****-/-*** mice. Data are mean ± SE. n=5-7 *p<0.05, **p<0.01, ***p<0.001, ****p<0.0001.

As compared to WT mice, *Cd36^-/-^* mice had higher glucose uptake (Rg) in leg skeletal muscle (superficial vastus lateralis and soleus) as well as in brown and subcutaneous white adipose tissues. Glucose uptake by the heart and gastrocnemius trended higher but did not reach significance (**Fig. 1G, H**). The hyperinsulinemic glucose clamp data support the interpretation that CD36 deletion improves insulin-stimulated glucose disposal through increases in Rd and in insulin suppression of endogenous Ra.

### Muscle insulin signaling is enhanced in Cd36^-/-^ mice

The enhanced insulin stimulated systemic glucose disposal observed in *Cd36****^-/-^*** mice (**Figure 1**) is not recapitulated in mice with muscle or cardiomyocyte specific CD36 deletion, which associate with reduced expression of insulin signaling and glucose metabolism genes in muscle and heart (5, 16). CD36 knockdown in cultured muscle cells also suppresses insulin action to activate AKT by removing CD36 enhancement of insulin receptor phosphorylation and signaling (5). We examined insulin activation of AKT in muscle of *Cd36****^-/-^*** mice to determine if it correlates with the enhanced glucose disappearance observed in these mice during the insulin clamp. First, we confirmed as shown earlier (15) that *Cd36****^-/-^*** mice as compared to wildtype (WT) controls have enhanced glucose tolerance and are protected from glucose intolerance induced by a high fat diet (HFD; 60% fat and 7% sucrose) (**Fig 2A, B, C),** ruling out alterations in insulin action due to changes in mice environment or breeding. Insulin signaling was assessed in muscle of 5h fasted WT and *Cd36^-/-^* mice given 0.75 U/kg insulin or saline (controls) intraperitoneally (IP). The mixed fiber quadriceps muscle was harvested 15 min after insulin injection and tissue lysates probed for AKT, pAKT and CD36. Muscles of *Cd36****^-/-^*** mice have similar basal AKT^S473^ and AKT^T308^phosphorylation levels as WT mice but a greatly enhanced insulin-stimulated AKT phosphorylation (∼7 fold as compared to ∼3 fold in WT) with no change in total AKT (**Fig. 2D, E,F**) consistent with enhanced muscle insulin signaling and with the findings from the insulin clamp experiments.

**Figure 2.**
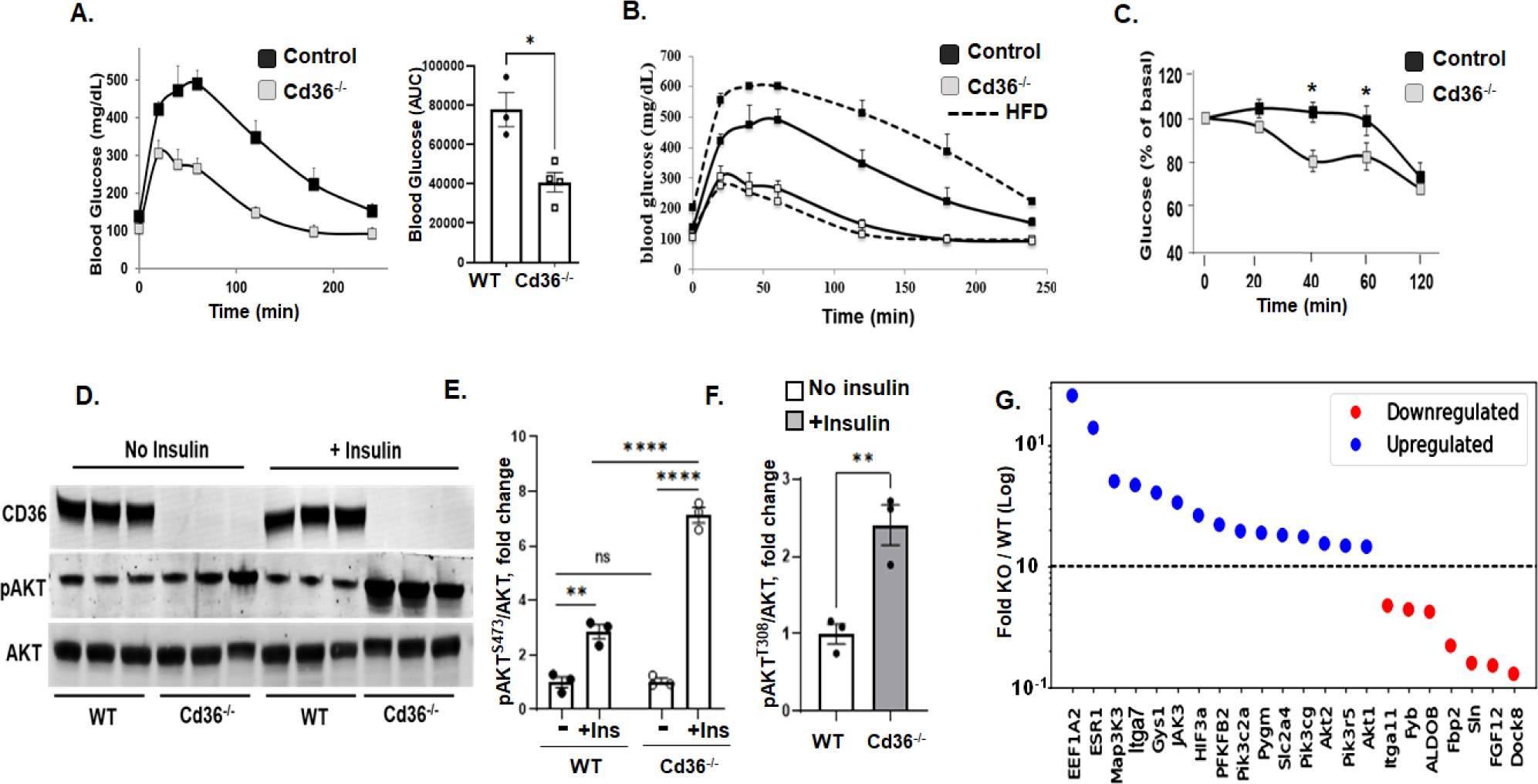
Enhanced Insulin signaling in muscle of *Cd36-/-* mice. **A.** Intraperitoneal (IP) glucose tolerance test (IPGTT) in 5h fasted mice showing enhanced glucose disposal in *Cd36****-/-*** as compared to WT mice. **B.** IPGTT, and **C.** Insulin sensitivity test in mice after feeding a high fat diet (HFD) showing glucose intolerance in WT mice, while *Cd36****-/-*** mice are protected. N=5-6 per group. **D.** 5h fasted *Cd36****-/-*** mice and WT controls were given insulin (7.5 IU/kg) IP and the gastrocnemius muscles were harvested 15 min later and processed for Western blots. Shown are signals for CD36, pAKT^S473^ and total AKT. **E**-**F.** Levels of insulin phosphorylated AKT (S473 and T308) adjusted for total AKT. **G**-**H.** Enhanced S6 phosphorylation in gastrocnemius of *Cd36****-/-*** mice as compared to WT mice and quantification of levels adjusted for total S6. **I .** Altered expression of genes related to insulin action and glucose metabolism in muscle of WT and *Cd36****-/-*** mice, n=3 per group. *p<0.05, **p<0.01, ****p<0.0001.

We previously reported in 5h fasted *Cd36****^-/-^*** mice significantly enhanced [¹⁸F]-Fluorodeoxyglucose (2-FDG) uptake by various muscles. The fold increase over WT was 5 for heart, 4 for soleus, 3 for diaphragm and gastrocnemius and 2 for hindlimb (15). Glycogen and triglyceride levels were reduced in heart and muscle. Examination of gene expression in hearts from 5h fasted mice showed upregulation of genes that function in insulin signaling or glucose utilization (**Fig. 2G**). Increased expression of Glut4 (Slc24, 1.8-fold), 6-phosphofructo-2-kinase/fructose-2,6-bisphosphatase (Pfkfb2), which regulates glycolytic flux (2-fold), so is expression of hypoxia inducing factor HIF3a (2.6-fold) which upregulates Pfkfb2 (30) and of glycogen synthase (Gys1, 4-fold) and Glycogen phosphorylase (Pygm,1.9-fold), which regulate glycogen turnover. Expression of PI3K, the pathway that mediates insulin effects on glucose metabolism (pik3c2a and pik3cg) was 1.7-fold higher, those of AKT2 (1.5-fold), AKT1 (1.45-fold) and AKT1 substrate 1 (2.3-fold). In addition, fibroblast growth factor receptor 1(FGFr1) a major target of FGF and potent stimulator of glucose metabolism in muscle (31) is increased 1.9-fold, insulin like growth factor receptor 1 (IGFR1), which binds IGF, and insulin is up 1.7-fold. Several integrins (ITG) are upregulated. Integrins interact with the extracellular matrix and maintain cell attachment, survival and growth by regulating the signaling of PI3K/AKT, growth factors (FGF, IGF) and Src kinases (32). In addition to a 4.7-fold upregulation of ITGa7 (**Figure 2G**), there were increases in expression of ITGa9, 4.7-fold, ITGa5, 3-fold, and ITGa3, 2.3-fold (data not shown). ITGA7/beta1 is the primary receptor for basement membrane laminin in skeletal and heart myofibers and is important in regulating myofiber survival (33, 34). Expression of Janus Kinase 3 (JAK3) which stimulates AKT-independent glucose uptake (35) increased 3-fold **Figure 2G**.

### *Cd36^-/-^* mice have normal transendothelial insulin flux but vascular dysfunction

#### Insulin Flux

Insulin access to muscle cells is regulated by the capillary endothelium, which limits the rate of glucose disposal (19, 20). We examined whether CD36 deletion might affect transendothelial insulin transport (EIT). To investigate this, we used the exposed gastrocnemius preparation and intravital imaging of fluorescent insulin-647 (INS-647) as described under methods and in greater detail previously (24). Plasma INS-647 dispersion to pericapillary interstitium was unaltered in *Cd36^-^*^/-^ mice. Plasma (**Fig. 3A)** and interstitial INS-647 (**Fig. 3B)** was similar for *Cd36^-^*^/-^ and WT mice. The decrease in ratio of plasma INS-647/interstitial INS-647, which measures rate of transfer of capillary insulin with interstitial insulin was identical in both groups (**Fig. 3C**) with gradient decay constants of ∼0.15 min^-1^ (**Fig. 3D**). These data indicate that transendothelial insulin flux is not altered in *Cd36****^-/-^*** mice and did not contribute to the enhanced insulin stimulated glucose disposal.

**Figure 3.**
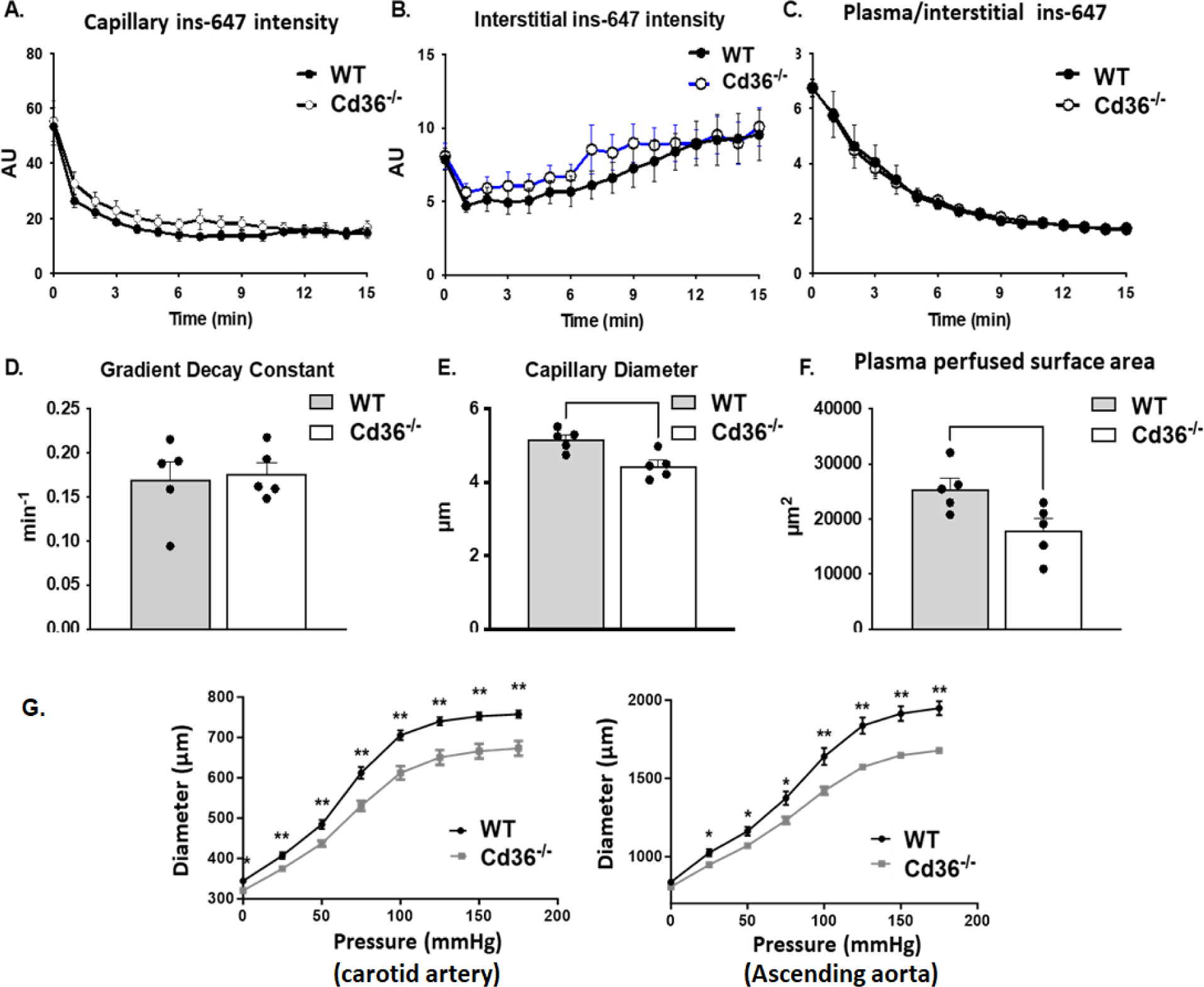
Trans-endothelial insulin efflux in muscle and the microvascular perfused area. WT and *Cd36-/-* mice were given a 2 U/kg bolus of insulin-647. **A.** Capillary plasma INS-647 fluorescence **B.** INS-647 in interstitial space extending 1-3 µm from capillary wall. **C.** Plasma/interstitial INS-647 intensity. **D.** Exponential decay constant of plasma/interstitial INS-647 gradient. **E.** Capillary diameter and **F**. Perfused capillary surface, both assessed with 2 MDa tetramethylrhodamine-dextran fluorescence, a vascular marker, show ∼30% reduction in *Cd36-/-* mice compared to WT mice. Data are means ± SE. n=5 per group. **G.** Intravascular pressure of carotid artery (left panel) and aorta (right panel) from *Cd36****-/-*** mice compared to WT mice (n=3/genotype). The vessels were mounted in saline on a pressure arteriograph, pressurized and longitudinally stretched three times to *in vivo* length before data capture. *p<0.05, **p<0.01.

#### Capillary diameter and perfused area

Fluorescence of rhodamine-dextran was used to determine the capillary diameter (distance between glycocalyx on opposing endothelium) and the perfused capillary cross sectional surface area. Capillary diameter (**Fig. 3E**) and the perfused capillary surface area (**Fig. 3F**) are reduced by ∼15 and ∼30%, respectively, in *Cd36****^-/-^*** as compared to WT mice. The decrease in perfused capillary surface area is unexpected considering the increased insulin action in *Cd36^-^*^/-^ mice. To confirm this finding, we examined changes in microvascular perfusion in *Cd36^-^*^/-^ mice using another organ and a different method. Arterial Spin Labeling Magnetic Resonance Imaging (ASL-MRI), a non-invasive, quantitative measure of microvascular tissue perfusion (36) was used to estimate renal perfusion. The ASL-MRI data (n=5 mice/group) collected at baseline under anesthesia (1% isoflurane in 100% O2) using a flow-sensitive (FAIR)-type ASL-MRI protocol (37) revealed a 34% deficit (p<0.05) in capillary renal perfusion in *Cd36****^-^*^/-^** mice (p<0.05) (data not shown) confirming the observation (**Fig. 3F)**.

#### Vessel compliance

The increase in diameter of the carotid and aorta arteries in response to increased intravascular pressure was examined for *Cd36****^-/-^*** mice and WT controls. Vessels from *Cd36****^-/-^*** mice as compared to those from WT mice showed reduced dilation as intravascular pressure increased from 0 to 175 mmHg in 25-mmHg steps (**Fig. 3G**). Thus, reduced vessel compliance might have contributed to measurements of smaller capillary diameter and reduced capillary perfused surface area in *Cd36****^-^*^/-^** mice.

In summary insulin action on glucose uptake by muscle is enhanced in *Cd36****^-^*^/-^** mice but this enhancement is independent of microvascular adaptations that improve insulin delivery. Transcapillary flux of insulin is normal in *Cd36****^-/-^*** mice, however there is evidence of endothelial dysfunction manifested by a diminished perfused capillary area and reduced vessel compliance. Although endothelial dysfunction normally associates with diminished glucose disposal, paradoxically in the case of CD36 deficiency it coexists with a higher insulin stimulated glucose disposal.

### Individuals with partial CD36 deficiency have enhanced insulin response of glucose disposal but microvascular insulin resistance

Common SNPs that reduce CD36 protein have been identified in many populations including African Americans (38) and Caucasians (10). Carriers of the G-allele of coding SNP 3211938 have 50% of normal CD36 levels (9, 38). We previously reported on presence of endothelial dysfunction in these individuals (9) but there is no information on whether this associates with improved or compromised insulin sensitivity.

#### Demographic Characteristics of subjects

Thirty-five subjects were screened, fourteen were excluded for not meeting inclusion criteria or withdrew consent, and twenty-one completed the study. Participants were divided into two groups based on the CD36 rs3211938 genotype. Controls (n=13) were homozygous for the major allele (T/T) and have normal CD36 expression, whereas carriers (n=8) of the minor allele G (G/T) have ∼50% reduced CD36 expression (9, 10, 38). Only one subject was homozygous (G/G) with 100% reduced CD36 expression and was included with the G/T cohort. The G-allele carriers were on average 8 year older than non-carriers and their average weight trended lower. Fasting glucose and triglyceride levels were similar. No participants were hypertensive; systolic (SBP) and diastolic blood pressure (DBP) were similar. All participants were healthy and not taking medications except for birth control in women (**Table 1**).

**Table 1.** Demographic characteristics of study participants.

|  | <u>G-allele</u> N=8 | <u>Non-carriers</u> N=13 |  |
| --- | --- | --- | --- |
| Age, years | 42 ± 2.6 | 34 ± 2.1 | 0.019* |
| Gender(m/f) | 3/5 | 3/10 |  |
| Weight, kg | 79 ± 2.9 | 89 ± 3.8 | 0.083 |
| Glucose, mg/dL | 81 ± 3.8 | 86 ± 5.4 | 0.511 |
| Cholesterol, mg/dL | 180 ± 13.1 | 177 ± 6.7 | 0.821 |
| HDL, mg/dL | 51 ± 4.88 | 54 ± 3.5 | 0.567 |
| LDL, mg/dL | 106 ± 12.8 | 106 ± 6.1 | 0.986 |
| Triglycerides, mg/dL | 116 ± 26.1 | 83 ± 10.9 | 0.189 |
| SBP, mm Hg | 124 ± 4.7 | 120 ± 2.4 | 0.349 |
| DBP, mm Hg | 75 ± 3.6 | 77 ± 2.5 | 0.857 |
| HR, bpm | 78 ± 7.5 | 68 ± 3.7 | 0.206 |

#### Insulin fails to increase microvascular blood volume in G-allele carriers

We examined insulin recruitment of the microvasculature in rs3211938 G-allele carriers (G/T) and control non-carriers (T/T). In non-carriers insulin increased microvascular blood volume index (MBVi) from 8.4±0.63 to 11.3±1.37 (35%, p=0.05). In contrast, no insulin-induced increase occurred in G-allele carriers (G/T) (**Fig. 4A**). Intralipid infusion instead of saline before the hyperinsulinemic clamp blunted insulin’s effect on microvascular recruitment in non-carriers and G-carriers remained unresponsive (**Fig. 4B)**.

**Figure 4.**
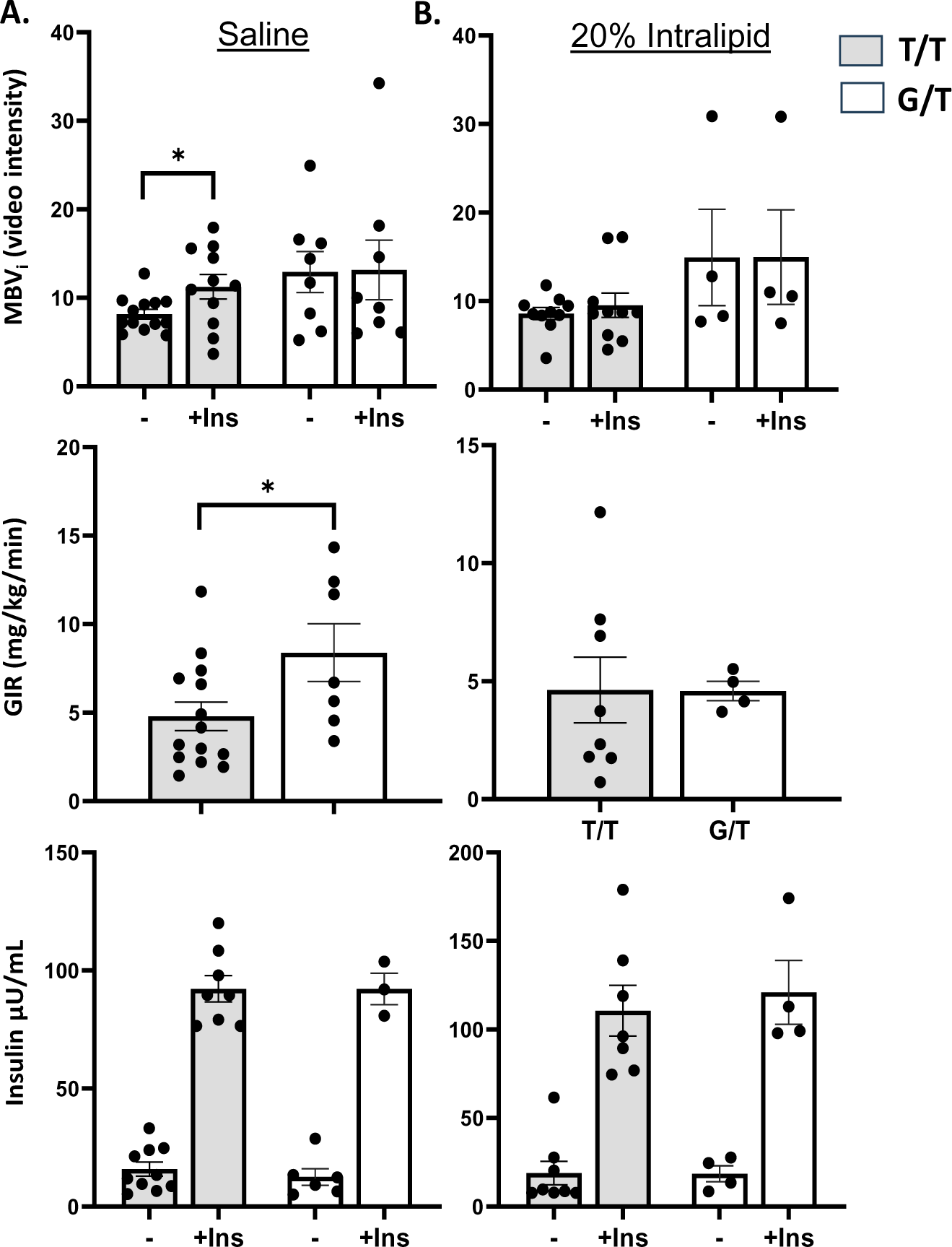
Hyperinsulinemic euglycemic clamp in individuals carrying the G allele (G/T) of CD36 SNP rs3211938 with partial CD36 deficiency versus non-carriers controls (T/T). Participants received standard saline infusion (45ml/h) for 6 h, and a hyperinsulinemic-euglycemic (HIE) clamp during the last three h. Insulin was infused at a rate of 80 mU·m-2·min-1 for 5 min followed by 40 mU·m-2·min-1 for the rest of the study. A. Upper panel: Effect of insulin on microvascular blood volume index (MBVi). Middle panel: Glucose infusion rate (GIR). Bottom panel: Circulating insulin levels were similar for G- allele carriers and noncarriers during basal and clamp periods. B. Participants received a 6-hour IV infusion of 20% Intralipid before the three-hour HIE. Upper panel: MVBi, middle panel: GIR, bottom panel: circulating insulin levels. *P=0.05

#### Enhanced whole body insulin sensitivity in G-allele carriers versus non-carriers

CD36 SNPs have been linked to the metabolic syndrome and T2D, but there is no definitive data on whether CD36 deficient subjects are insulin sensitive or resistant. We determined the glucose infusion rate (GIR) in both groups (G-allele carriers and noncarriers) during a continuous insulin infusion at a dose of 40 mU/m^2^/min. The G-allele carriers required 1.9-fold higher GIR than non-carriers, indicating they were more insulin sensitive **(Fig. 4C)**. Intralipid infusion decreased GIR in both carriers and non-carriers and the G-allele carriers were no longer more insulin sensitive as compared to non-carriers (**Fig. 4D**). The increased GIR in G-allele carriers during saline infusion did not reflect differences in insulin levels during the steady state phase of the clamp (**Fig. 4E**) and similar insulin concentrations were also measured during the intralipid infusion (**Fig. 4F)**.

In summary the insulin clamp findings indicate enhanced GIR supporting better insulin stimulated glucose utilization in G-allele carriers. This enhancement occurs without insulin stimulation of MVB flow, which suggests insulin resistance of the microvasculature in individuals with partial CD36 deficiency.

### CD36 knockdown in human microvascular cells impairs insulin action and eNOS activation

Signaling by insulin contributes to active maintenance of blood vessels through regulated production of NO by endothelial nitric oxide synthase (eNOS) and polymorphisms in eNOS associate with insulin resistance and T2D (39). To gain insight into the mechanism of the microvascular insulin resistance in G-allele carriers, we investigated role of CD36 in insulin regulation of eNOS using primary human-derived microvascular endothelial cells, hMEC, which express high levels of CD36, a microvascular signature gene (28, 40).

#### CD36 depletion in hMEC influences gene expression and insulin action

Cells were treated with either control siRNA or anti CD36 siRNA. First, the resulting alterations in gene expression were determined by RNA seq (**Fig. 5A**). CD36 depletion associated with increased expression of FGF1, insulin like growth factor 1 (IGF1), Jak3, PIK3 catalytic domain and various integrins (ITG) including ITGA11, ITGA4, ITGA5 and ITGB5. Integrins regulate signaling by PI3K and growth factors (32). Overall, the above upregulated genes function to promote glucose utilization and cell survival, either directly or through activation of PI3K/AKT and growth factor signaling (32, 35).

**Figure 5.**
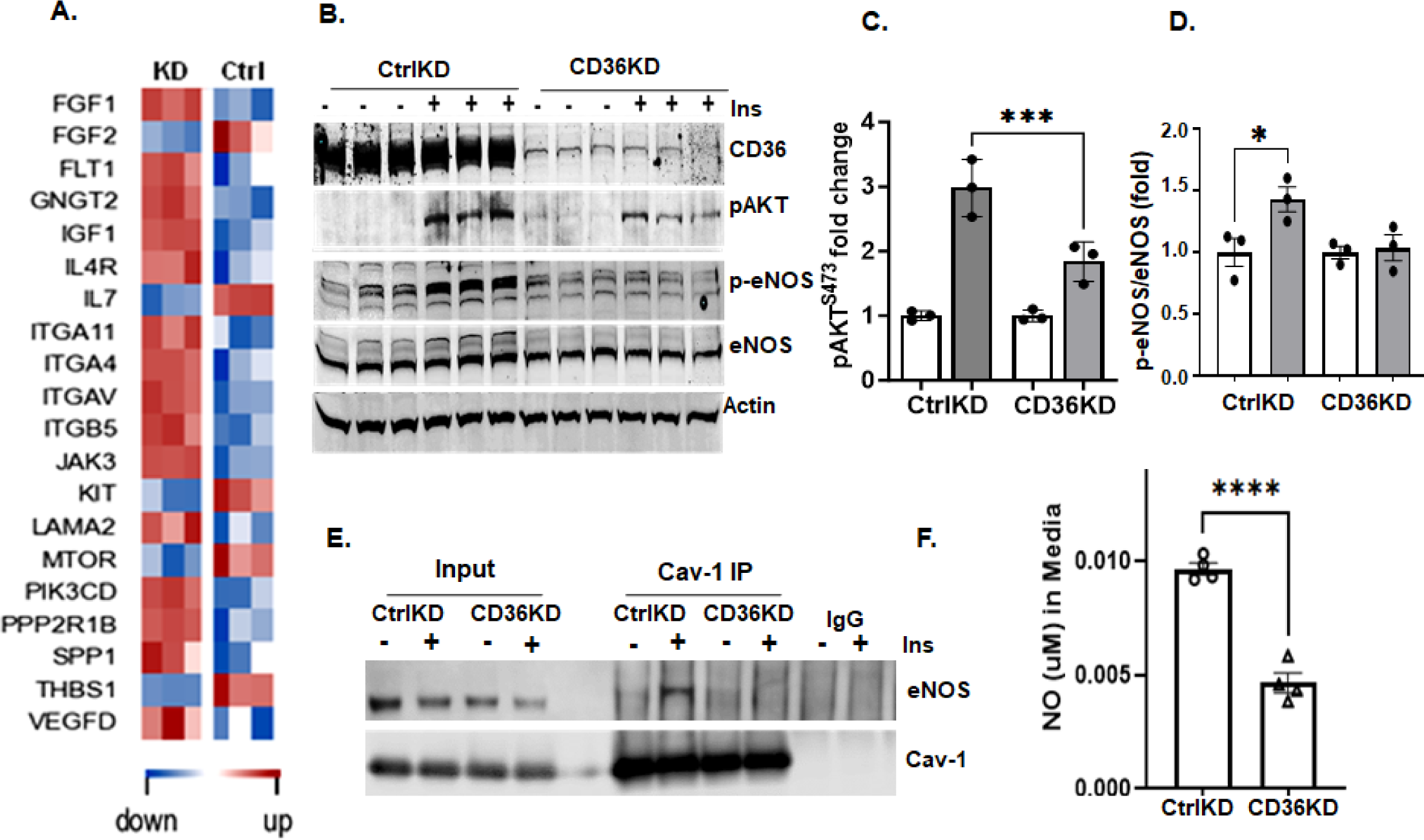
CD36 regulates insulin signaling in primary derived human microvascular cells (hMEC). **A**. Alterations in genes related to PI3K-Insulin signaling in control hMEC (Ctrl, treated with scrambled siRNA) and CD36KD (KD, hMEC treated with anti-CD36 siRNA). **B.** Western blots of hMEC, Control (CtrlKD) and CD36KD showing effect of insulin (100nM, 5min) on phosphorylation of AKT and eNOS. **C and D.** Densitometry analysis of pAKT/total AKT and of p-eNOS/total eNOS (n=3 p<0.05). **E**. CD36 knockdown on insulin- dependent interaction of Cav-1 and eNOS. Western blot is representative of three independent experiments. **F.** CD36 regulation of NO production. Ctrl and CD36KD hMEC were serum starved and subjected to insulin stimulation (100nM, 5min). n=4, assays. *p<0.05, ****p<0.005.

We previously showed that CD36 expression in human derived primary muscle cells facilitates IR signaling and its depletion suppresses insulin activation of AKT (5). The hMEC were tested for insulin responsiveness and the effect of CD36 knockdown. The hMEC behaved like the human muscle cells; they responded to insulin with robust phosphorylation of the activating sites of AKT^S473^ and eNOS^S1177^, and CD36 knockdown significantly suppressed insulin activation of both proteins (**Fig. 5B, C, D**).

#### Interaction of eNOS with Caveolin-1

In endothelial cells, Cav-1 interaction with eNOS regulates eNOS activity and this interaction is enhanced by insulin (41, 42). CD36 resides in caveolae and can influence Cav-1 dynamics (28) so we examined whether CD36 it influences insulin regulation of the Cav-1/eNOS interaction. Immunoprecipitation of Cav-1 from control and CD36 depleted hMEC showed, consistent with previous findings, that insulin enhances interaction of Cav-1 with eNOS in control cells (41). Interestingly, CD36 depletion eliminated the effect of insulin on Cav-1/eNOS interaction (**Fig. 5E**). In addition, consistent with CD36 knockdown acting to suppress insulin activation of eNOS, insulin-stimulated NO production was simultaneously reduced **(Fig. 5F**).

Together these data support influence of endothelial CD36 on insulin-induced eNOS activation and NO production and suggest that CD36 depletion associates with endothelial insulin resistance and reduced NO availability. These data are consistent with the lack of insulin induced increase in microvascular volume that is observed in G-allele carriers by the hyperinsulinemic clamps (**Fig. 4A**) and the defect in arterial flow mediated dilation and low cGMP levels previously described in these individuals (9). They are also consistent with the observed vascular dysfunction; reduced capillary perfusion and vessel compliance, in *Cd36****^-/-^*** mice (**Fig. 3** **E, F, G**).

Defects in microvascular insulin action and NO production and reduced endothelial compliance are a hallmark of metabolic disease and thought to be involved in etiology of T2D (43, 44). Previously we applied Predixcan analysis to a large patient sample in the BioVu database of Vanderbilt University and documented that genetically determined low CD36 mRNA in muscle or heart strongly associates with incidence of type 2 diabetes (T2D) and its complications (5). PrediXcan estimates for GWAS samples the Genetically Regulated eXpression (GReX) of a gene in specific tissues and uses GReX to identify genes associated with disease risk (45, 46). We again used PrediXcan to test whether the association holds if the analysis considers CD36 mRNA in blood or arteries. We found that low vascular CD36 levels associates with T2D and its renal, ophthalmic, and neurological manifestations (**Table 2**).

**Table 2:** Association of genetically determined low endothelial CD36 expression with T2D status and complications.

| Tissue CD36 | Effect | p-value | T2D and complications |
| --- | --- | --- | --- |
| Whole Blood | -4.323 | 0.000015 | T2D + renal manifestations |
| Artery -Tibial | -4.058 | 0.00005 | T2D + renal manifestations |
| Artery -Tibial | -3.506 | 0.00045 | T2D + ophthalmic manifestations |
| Artery -Tibial | -3.434 | 0.00059 | T2D + neurological manifestations |

CD36 contribution to T2D risk was evaluated using PrediXcan analysis (45). The genetically determined component of CD36 expression was estimated from gene expression imputation trained with reference transcriptome data (N = 44 tissues, 449 donors, version 6p) from the Genotype-Tissue Expression (GTEx) Consortium. Analysis was applied to 4702 total patients of European ancestry with 1484 patients with T2D, in the BioVU database (58) of Vanderbilt University where health records are tied to a DNA biobank. Samples were genotyped, Illumina 660K, and genotype imputation used 1000 Genomes Project (59) as reference. For replication of CD36 gene level association with T2D risk, we used GWAS meta-analysis data on HOMA-IR from the MAGIC Consortium (60) and applied PrediXcan to GWAS summary statistics. The replication data set had 37037 participants of European ancestry.

## Discussion

The coding CD36 SNP rs3211938 (T/G) is the major cause of CD36 deficiency in African Americans with an incidence of 9-12% for homozygous (G/G) and 20-25% for heterozygous (G/T) (9, 10). In Japanese, CD36 deficiency has a frequency of 3% for homozygous and 6% for heterozygous (14). Total deficiency is low in Caucasians (0.3%) but SNPs that reduce CD36 expression are relatively common (5-40%) (6, 11). CD36 SNPs often associate with high circulating lipids, FA (6, 9) and chylomicron remnants (11, 47) and with risk of metabolic syndrome (10, 48) and T2D coronary heart disease (6). We identified in the BioVu patient biobank association of low heart and muscle CD36 mRNA with T2D renal, ophthalmic and neurologic complications (5) and now confirm this association for low blood and vascular CD36 (**Table 2**). Mechanisms underlying CD36 link to T2D are largely unclear.

There is currently no data to support a link between CD36 deficiency and insulin resistance of glucose metabolism. A few studies in Japanese with CD36 deficiency documented blood lipid abnormalities but provided no definitive answer related to presence of insulin resistant glucose disposal (12, 49). In this study, our hyperinsulinemic euglycemic clamps in individuals with partial CD36 deficiency (G-allele carriers) (9) and matched controls document enhanced insulin stimulated glucose disposal in the CD36 deficient group, like what is observed in CD36 deficient mice. The enhanced insulin stimulated glucose disposal in G-allele carriers occurs without the insulin induced increase in microvascular blood volume. The MVB increase which normally enhances insulin delivery to muscle (19, 20) does not play a role in the improved insulin stimulated glucose disposal of G-allele carriers. The lack of insulin MVB response suggests presence of selective microvascular insulin resistance in G-allele carriers as compared to non-carriers (**Fig. 4A**). This finding is consistent with the previous demonstration of impaired flow mediated dilation of the brachial artery and low cGMP levels in these individuals, suggesting low NO bioavailability (9). Our data with hMEC (**Fig. 5**) directly link insulin resistance to endothelial dysfunction of the microvasculature. CD36 knockdown, blocks insulin signaling to AKT and insulin activation of eNOS and NO release. This provides a plausible mechanism for the endothelial dysfunction present in humans with partial CD36 deficiency and would apply to the reduced capillary perfusion and vessel compliance observed in *Cd36****^-/-^*** mice. Importantly in CD36 deficiency the microvascular dysfunction did not cause insulin resistance of glucose metabolism as would have been expected and as previously shown in obese mice (50). The major difference in the case of CD36 deficiency is that it reduces FA uptake and is critical endothelial FA transcytosis (16, 28). Infusion of intralipid in our study blunted the effect of insulin to increase MVB in non-carriers and reduced GIR in both groups (**Fig. 3B, C, D**) indicating that partial CD36 deficiency does not protect against excess intake. CD36 deficiency protects from the muscle insulin resistance induced by high fat feeding (**Fig. 2**) (15). The lack of protection in humans could reflect the difference between effects of total (mice) versus partial CD36 deficiency (G-allele carriers). However, uptake of FA from chylomicrons is not mediated by CD36 (51, 52) and the same is likely to apply to lipid emulsions infused along with heparin. In both cases lipolysis results in excessively high FA levels at the endothelial interface and under such conditions FA uptake is independent of CD36.

Our studies in *Cd36****^-/-^*** mice provide data that parallel and support interpretation of the findings in CD36 deficient humans. Enhanced insulin stimulated glucose uptake by heart, various muscles and adipose tissues, and better suppression of endogenous glucose production by the liver are observed in *Cd36****^-/-^*** mice as compared to controls (**Figs. 1, 2**). The enhancement in insulin action on glucose metabolism in muscle, liver and adipose tissue of *Cd36****^-/-^*** mice occurs despite a reduced capillary perfusion area and impaired vessel compliance in these mice. The mice studies, like the human studies indicate that enhanced capillary insulin delivery does not contribute to the improvement in insulin stimulated glucose disposal. The data in mice suggest an important role for the adaptive transcriptional remodeling of muscle in CD36 deficiency, where there is upregulated expression of GLUT4, of PI3K/AKT and of enzymes of glucose utilization. In addition, the primary receptors for two potent stimulators of muscle glucose uptake FGF-R1 and IGF-R1 are upregulated. Interestingly, expression of the corresponding growth factors FGF1 and IGF1 is increased in hMEC after CD36 knockdown, as compared to the low levels in cells treated with control siRNA. CD36 antagonizes effects of growth and pro-angiogenic factors in EC (53). As its deletion relieves this negative regulation, one can speculate that this increases endothelial release of FGF1 and IGF-1 by CD36 deficient EC. Interaction of the growth factors with their receptors in muscle would enhance glucose uptake and utilization.

The endothelium can regulate insulin-stimulated glucose disposal through controlling insulin delivery to muscle (20), through release of NO (54, 55) and other bioactive molecules (56) or the secretion of exosomes (28). Endothelial insulin resistance is thought to be a precursor of muscle insulin resistance and the reduced NO availability was implicated in driving insulin resistance of glucose metabolism in obese mice (50). However, an opposite outcome like the one we describe in G-allele carriers was reported in a recent study where inducing EC specific insulin resistance by disabling endothelial IR and IGF-1R, enhanced muscle and fat insulin sensitivity (57). Increased expression of NADPH oxidase 4 and consequent endothelial release of peroxide were suggested as mediators. We do not observe increased NADPH oxidase 4 expression in CD36 deficient EC, although we do not rule out its potential contribution. In the case of CD36 deficiency we believe the effect might be linked to the absence of endothelial FA uptake. Endothelial cell CD36 controls delivery of circulating FA to heart and muscle (16) through a transfer mechanism that involves generation and secretion of CD36-containing FA-exosomes.

These EC derived exosomes can regulate muscle gene expression and metabolic adaptation (28). The exosome cargo and how it is altered in obesity or with excess fat intake will need to be examined as this might shed more light on the role of the endothelium in regulating insulin resistance.

In conclusion partial CD36 deficiency in humans, as observed in CD36 deficient mice increases insulin-stimulated glucose utilization, which energetically compensates for the suppressed FA uptake. Some limitations apply to our hyperinsulinemic euglycemic studies in humans notably the relatively small number of subjects studied (13 controls and 8 G-allele carriers). However, study participants underwent in-depth metabolic phenotyping before matching the cohorts. The small difference in weight between the cohorts is another limitation but it was addressed through adjusting the glucose infusion rate by weight.

The evidence provided in this study support the role of CD36 deficiency in microvascular insulin resistance and reduced NO bioavailability (**Fig. 4 and 5**). Although this impairment does not associate with reduced muscle glucose disposal and instead enhances it, the long-term effects of this dysfunction within the context of obesity are likely to be detrimental and could play an important role in the etiology of T2D complications. Genetic data associate low CD36 mRNA in various tissues including heart, skeletal muscle (5) and endothelium (**Table 2**) with T2D renal, ophthalmic and neurologic complications, which often reflect vascular impairments. A common CD36 haplotype in Caucasians associates with coronary heart disease in diabetics (6). The defect in NO production in CD36 deficiency identified in both CD36 deficient mice and humans is likely to contribute to etiology of the microvascular complications of T2D and could be targeted to prevent or mitigate these abnormalities.

## Data Availability

All data produced in the present study are available upon reasonable request to the authors

## Acknowledgments

We acknowledge support of National Institute of Health RO1DK111175 (NAA, CAS, NNA), NHLBI R01HL157584 (CAS), NHLBI R01 HL045095 (IJG, NAA) and Clinical Translational Science Award (CTSA) 5UL1TR002243-03 (CAS), and assistance of Vanderbilt Institute for Clinical and Translational Sciences and Washington University Nutrition and Obesity Center (NORC, NIH P30 DK056341).

